# Self-Rehabilitation Strategy for Rural Community-dwelling Stroke Survivors in a Lower-Middle Income Country: A Delphi Study

**DOI:** 10.1101/2024.05.03.24306686

**Authors:** Rabi’u Ibrahim, Conran Joseph, Aimée Stewart, Isa Usman Lawal

**Author notes:** Corresponding author (RI). These authors contributed equally to this work (Rabi’u Ibrahim, Isa Usman Lawal). These authors also contributed equally to this work (Conran Joseph, Aimée Stewart).

## Abstract

**Background:** More than half of stroke survivors in LMICs lack access to stroke rehabilitation services. The promotion of self-rehabilitation could be promising in an attempt to address the stroke rehabilitation inadequacies in LMICs. Self-rehabilitation interventions can easily be accepted by many community-dwelling stroke survivors, and therefore, have the potential to drive towards the successful realization of Sustainable Development Goals and other WHO rehabilitation goals. We report a consensus building process that sought to identify which task training is relevant to include in a task-specific self-rehabilitation strategy for the rural community-dwelling stroke survivors.

**Methods:** We used an iterative two-staged mixed-method consensus-building approach: (1) focus group discussions (n = 5) with rural community-dwelling stroke survivors were conducted to explore personal life experiences in performing daily activities, and the results were used to developed a lists of candidate task trainings that could be included in a task-specific self-rehabilitation intervention model for improving functional ability post-stroke survivors; (2) a three-round Delphi exercise with a panel of stroke rehabilitation experts to establish consensus on the importance/relevance of the developed task trainings. Consensus was pre-defined to be the point where the proportion of items giving a rating of 3 (quite relevant) or 4 (highly relevant) by raters would be ≥ 0.8.

**Results:** A list of 74 task training was generated from the results of the focus groups and grouped as follows: training for the upper extremity (37); lower extremity training (21); while 7 and 9 task trainings were grouped under the trunk and balance training respectively. A panel of 13 experts in the Delphi reviewed these task trainings and consensus was achieved on keeping 28 task trainings in the first round and an additional 7 in the second round. In the study team’s analysis of open text responses, several areas of debate were identified and some task trainings were modified. The exercise yielded 49 trainings (66% of 74) on which there was consensus (the mean proportion of items giving a rating of 3 or 4 by raters was 0.93) to keep 3 task training groups relating to: upper extremity (27), lower extremity/balance (8), trunk strength (4) and warm up exercises (10).

**Conclusions:** The study provides a consensus-based view of the features of a task-specific self-rehabilitation training strategy to improve outcomes following a stroke. This self-rehabilitation training strategy can be used as an intervention approach to augment and promote stroke rehabilitation among the rural community-dwelling stroke survivors, especially in SSA.

## Introduction

Stroke is a substantial source of acquired adult neurological disability worldwide.^1^ In low- and middle-income countries (LMICs), stroke incidence has increased alarmingly in the past decade.^2,3^ The consequences of stroke have long-standing effects and require long-term management of the ensuing limitations. Lack of resources, inadequate numbers of rehabilitation professionals, poor awareness, and lack of technical capacity have made the accessibility and availability of stroke rehabilitation services difficult in LMICs, particularly the rural areas of sub-Saharan Africa (SSA).^4^

Currently, the needs for stroke rehabilitation are largely unmet. In some regions of LMICs, more than 50% of people do not receive the rehabilitation services they require.^5^ Disturbances including conflicts, disasters and outbreak of infectious diseases create massive surges in rehabilitation needs while also disrupting any available rehabilitation services. The effect of the recent COVID-19 pandemic and its associated physical distancing protocols, and other emerging communicable diseases add up to already existing challenges to stroke rehabilitation delivery in many regions of LMICs.

In Nigeria, like in other parts of LMICs, there is no existing intervention strategy to support rehabilitation among rural community-dwelling stroke survivors. Rehabilitation professionals employ the practice of prescribing certain activities termed as ‘home program’ for stroke survivors to run-through at home. These activities are not always task-specific and none has been established through a proper expert consensus approach.

The promotion of self-rehabilitation that include task-specific training could be promising in an attempt to address the stroke rehabilitation inadequacies in LMICs. Self-rehabilitation interventions have been shown to improve outcomes post-stroke, lessen the risk of stroke relapse and have encouraging impacts on healthcare resource utilization, which is of great implication for LIMICs.^6,7,8^ Moreover, having the advantage of being run-through in the comfort of an individual’s home, self-rehabilitation interventions can easily be accepted by many community-dwelling stroke survivors, and therefore, have the potential to promote wider stroke rehabilitation coverage and drive towards the successful realization of SDG goal number 3 - “Ensuring healthy lives and promoting well-being for all”^9^ and other WHO rehabilitation goals.^10^

To develop a structured task-specific self-rehabilitation strategy for community-dwelling stroke survivors, a Delphi approach would be more appropriate in order to achieve relevant experts’ consensus on the task trainings to be included in the rehabilitation strategy, also the stroke survivors should be consulted in developing the initial task items of the model. In research generally the Delphi method is used to build consensus around a particular research question or topic. Experts are surveyed via questionnaires without being physically assembled.^11^ A Delphi method is usually used in healthcare when guidelines or treatment protocols need to be developed, and when evidence is limited or inconsistent.^12^

The primary objective of this study was to build experts consensus on the appropriate item (task training) to be included in a task-specific self-rehabilitation training strategy that could be administered among Hausa-speaking community-dwelling stroke survivors in Nigeria.

## Methods

Ethical approval for this study was sought and obtained from the College Health Research Ethics Committee of Bayero University Kano (ref: NHREC/06/12/19/5). The ACCORD (Accurate Consensus Reporting Document) guideline^13^ was followed in reporting of this study.

A two-stage consensus-building approach was adopted in this study; the first stage was the identification of the relevant tasks that could be included on the basis of importance, in a task-specific self-rehabilitation intervention model for improving functional ability among community-dwelling stroke survivors in a resource-limited setting (September-December, 2022). The second stage comprised the conduct of a modified Delphi exercise to build consensus on the final list of task trainings to include in the intervention model (January 2023-January 2024).

### Stage 1: Identifying the task training items to include in the intervention model

A list of potential task trainings that could be entered into the intervention model was drawn from information gathered through focus group discussions with stroke survivors. Focus group discussion was used in a previous study to inform the initial content of a Delphi study instrument.^14^

#### Focus Group Discussions (FGDs)

Five FGDs involving twenty-nine community-dwelling stroke survivors were conducted (between 29th September to 3rd December, 2022), with the aim of identifying the common daily activities that stroke survivors find difficult to accomplish. The inhabitants of the study area are predominantly from Hausa tribe engaged in rice farming.

#### FGD: Participants and Sampling Strategy

In order to generate a diverse information with a range of experiences in performing daily activities after stroke, participants were purposefully selected based on gender blend, age, and in diverse locations within the community. Local community contacts and some identified stroke survivors within the community help with participants’ recruitment. Consent package which provided details of the study process was fully explained to the prospective participants and their written consent to participate was sought and obtained. All stroke survivors who expressed interest and met the following criteria: aged 18 and above years; diagnosed with ischemic or hemorrhagic stroke; speak and understand the Hausa language; capable of giving informed consent were recruited into the study. Participants were excluded if they have cerebrovascular events due to malignancy or head trauma; have limited comprehension (receptive and/or expressive aphasia); have been diagnosed with other neurological/mental disorder.

#### FGDs: procedure

FGD sessions were scheduled with those who gave consent and met the inclusion criteria. Four of the focus groups included 6 participants while one focus group included 5 participants. The discussions took place in a convenient classroom in one of the schools within the Kura town. Using a semi-structured interview guide (S1) which was developed by two members of the research team (IUL and RI), the participants’ experiences in performing daily activities after stroke were explored.

Discussions were conducted in Hausa language and audio-recorded. Maximum duration of each FGD was 1h and 40 min. The FGDs were facilitated by IUL and field notes were taken by RI. The concept of data saturation and collection of rich and thick data^15^ was used to end the data collection.

#### FGD: Analysis and results

The audio-recordings were transcribed verbatim and anonymized with codes. The Hausa language transcript was translated into English language at the English language department of Bayero University Kano. Using deductive open coding process, two members of the research team (IUL and RI) agreed on an initial flat coding frame, developed a set of predetermined codes based on the study’s objective and both read through the data line-by-line and assigned excerpts to codes until themes were developed.

Three major themes and eight sub-themes were identified (table 1). Themes were related to the most commonly reported activities/tasks which stroke survivors struggled to perform and include personal care activities, religious activities and daily participation activities.

**Table 1.** Themes and sub-themes.

| Themes | Sub-themes |
| --- | --- |
| Personal care | Feeding<br>Dressing/undressing<br>Washing<br>Bathing |
| Religious activities | Ablution<br>Praying |
| Participation | Standing/walking<br>Cooking |

These themes are described below with example of some anonymized quotes.

##### Personal care

The major activities reported under this theme are categorized into feeding, dressing/undressing and bathing and washing:

> “..I have no option but to eat with the left hand… I don’t like it, but I have to do it.” (P6). “I eat and drink with the left hand. I use a spoon to eat anything.” (P11).
>
> “..Now I use a walking stick to walk to the bathroom…My wife bathes me and wears my clothes for me” (P8).
>
> “Wearing trousers is the most difficult for me, so I first wear my shirt even in the bathroom. But I have to go to my bedroom and wear the trousers while seated by the bedside” (P16).
>
> “It is always difficult to tie a wrapper around my waist, because I use one hand, so my daughter has to assist me in tying it” (P12).
>
> “Whenever I enter the bathroom to bathe myself, I have to rely on the hand that is not affected, and in this manner I just bathe anyhow and come out” (P8).
>
> “..I try to bathe myself, but I have to do it slowly and I have to use the left hand to assist the right”. (P20).
>
> “to wash my clothes now is very difficult, and there is no one to assist me with that (P2)

##### Religious activities

The participants were predominantly Muslims and many of them reported having difficulty in performing ablution (sequential washing of some body parts which is a pre-requisite for performing the daily prayers) and praying:

> “When performing ablution, I have to sit down because I can’t squat down, the leg that supports half of my body is not well” (P7).
>
> “In the aspect of performing religious rituals, I am weak, thank God that now I can bend down during prayers, but I can’t prostrate. Also I perform ablution slowly, the person that assists me is also not feeling well now. So somehow I managed to do it, even though my hand is heavy” (P8).
>
> “..to perform ablution, someone have to assist me by pouring the water in to my unaffected hand, that’s how I do it” (P4).

##### Participation activities

Standing/walking and cooking activities were the main activities categorized under this theme:

> “If I am seated I have to use the right arm to stand up, but I do that slowly. But when I want to walk I have to hold onto something close by…when going out of my house, I have to use the wall as support, because I don’t use a walking stick. But honestly, I don’t go far as I am usually advised to” (P3).
>
> “..For example, when I want to stand up to walk, I do that from the right side of the body because it is the stronger side. Just not long ago I fell down while trying to get off a motorcycle, I tried getting down from the left side and place the left leg on the ground first, but it couldn’t carry me and gave way so I fell down…it was frustrating” (P21).
>
> “..So sometimes when I have to walk up or down a high surface, if I step up with the affected leg first, I stay still in that position, I cannot proceed. But when I step up with the unaffected leg first, then I can easily follow it with the left one and go up. But I can’t step down” (P9).
>
> “..You go out every day to work and earn a living for you and your family, but now I can’t. I rely totally on others to give me…I used to fry groundnuts and sell, but I can’t do that now because of the sickness, I am not able to cook” (P10).

#### Developing the initial task training items

This was done in two steps; first, the results of the focus group discussions were considered by three physiotherapists (IUL, RI and CJ) with more than 15 years’ experience in stroke rehabilitation. All the tasks that the stroke survivors reported as being difficult were identified. Secondly, the three physiotherapists in collaboration with a kinesiologist isolated and grouped similar tasks based on the pattern of movement and the body part (i.e. upper extremities, lower extremities and the trunk) involved. The training requirements in each group were further categorized based on the activities. This process resulted in the generation of a list of potential tasks (S2) to include in the intervention model which was used in the Delphi exercise.

### Stage 2: Consensus-building to select and refine the tasks to include in the intervention model

We used a modified Delphi technique^16^ to build consensus on which tasks generated from stage 1 were relevant to include in the intervention model. Three rounds of rating and review by an expert panel were conducted over an eleven-month period.

#### Selection of panelists

Using purposive sampling technique, thirty stroke rehabilitation experts (comprising of ten from Nigeria, ten from other parts of Africa and ten from other parts of the world) from diverse professional groups involved in stroke rehabilitation, with at least 15 years of experience in stroke management were contacted through email for their consent (S3) to participate in the Delphi process.

Potential panelists were identified through their published work in the field of stroke rehabilitation and some suggested by the main study team. These potential panelists were also asked to suggest others who they consider suitable to participate (snowball sampling), and were also invited, provided they met the inclusion criteria. Our aim was to achieve a panel of 11 to 30 panelists, which is a range of sample size regarded as effective and reliable for a Delphi technique.^17,18^ A copy of the set of tasks including an instruction note (S4) on how to rate the of items was sent to each of those who provided signed written consent. All correspondence with the panelists was done via e-mail.

#### Defining consensus

A consensus on a topic or an issue can be regarded as the general agreement among a group of people that are well informed (experts) about the topic or issue in question. In the literature, consensus was defined as the “gathering of individual evaluations around a median response with minimal divergence.”^19,20^ While experts can reach 100% agreement on only a few issues,^21^ however, consensus is determined by a certain percentage of experts who agree to an issue in a Delphi survey, and this should be defined and stated before the conduct of the survey.^22^

The level of consensus on what task training is relevant was measured at the end of first and second rounds in this study, while the final level of consensus on the overall suitability of the intervention model was measured and the end of the third round. The level of consensus on the relevance of each task training was determined as follows: for each task training, the number of experts giving a rating of 3 or 4 divided by the total number of experts was computed. Consensus was decided to be accomplished if a task training would receive a score of ≥ 0.78.^23^ Any task training that scored < 0.78 was either modified for the next round or removed. For the overall suitability of the intervention model, the level of consensus was determined as follows: The proportion of items giving a rating of 3 or 4 by raters involved was computed as advocated by Waltz and Bausell.^24^ A score of ≥ 0.8 would be accepted as moderate to high consensus based on available procedural evidence.^25,26,27,21^

#### Delphi round 1

In this round the panelists rate each item of the model in terms of its relevance to the underlying construct. The item ratings were on a 4-point scale to avoid a neutral and ambivalent midpoint^23^ and to produce stable findings in Delphi studies.^18^ The rating interpretation was as follows: 1 = not relevant, 2 = somewhat relevant, 3 = quite relevant and 4 = highly relevant. After all the panelists have finished the ratings, the package was retrieved and the computation to determine consensus on the item relevance was done. Items that were not rated by at least two-third of the panelists were not included in the computation and were regarded as items on which no consensus was reached and were included in the next round.

For each task item, options for the panelists to provide text comments in support of their rating, reason for abstaining from rating or to suggest changes to the task were available. The panelists were given a period of three weeks to respond and reminders were sent on a weekly basis.

#### Delphi round 2

In this round, the written opinion of all the panelist were summarized and shared among them. Experts were invited to consider their scores for the remaining task trainings that did not make the required item validity score and were not removed, based on group responses in relation to the overall responses received. They decided/suggested what they deemed appropriate between their rating and the average opinion of other respondents. Additionally, further explanation of some culture-based tasks was done to the panelists, particularly, those who were not familiar with the culture of the intervention target population. Panelists were also asked to give written opinions (S5) on the overall structure and inclusiveness of the included tasks. The item rating and validity score computation was as in round 1.

Prior to the commencement of the third round, an in-person meeting of the study team was held, purposely, to decide on some tasks which are peculiar to the culture of the study area and that some panelists were not familiar with, and as a result abstained from rating those tasks. The study team also, finalized the structure and inclusiveness of the task training items in the model by considering the comments from the panelists in round 2. All items that had the required score and no suggestions were made about them by the panelists, were not debated in the meeting and panel consensus was deemed final.

#### Delphi round 3

Round 3 involves the content validation of the task training items in the model (including those that have been retained in round 1 and 2, and those modified or added during the in-person study team meeting. In this round, panelists were invited to reassess the content and rate the whole model using the criteria in round 1. All responses were collected individually.

### Data analysis

Analysis of data was done using Microsoft Excel and involved calculating the proportions of agreement. For first and second rounds of the Delphi, the number of experts giving a rating of 3 or 4 (for each task training) divided by the total number of experts was computed. A score of ≥ 0.78 indicates consensus has been reached to keep the task training.^23^

For the third round of the Delphi, the average proportion of task training items giving a rating of 3 or 4 by raters involved was computed as advocated by Waltz and Bausell.^24^ Based on available procedural evidence, a content validity score of ≥ 0.8 was deemed consensus to keep the content of the intervention strategy.^25,26,27^

## Results

### Stage 1: Identifying the task training items to be included in the intervention model

Seventy-four (74) task training items (S2) were drawn and designed based on the information gathered through focus group discussions with stroke survivors, as potential items for the stroke self-rehabilitation model. Thirty-seven (37) task trainings were grouped as training for the upper extremity, 21 tasks were grouped under the lower extremity training, while 7 and 9 tasks were grouped under the trunk and balance training respectively. We further categorized the groups based on the activities involved (table 2).

**Table 2.** Initial Grouping/categorization and number of the task training items.

| **Task training group** | **Category** | **No of task training items** |
| --- | --- | --- |
| **Upper extremity training** | Trainings for reaching | 13 |
|  | Trainings for grasp/grip | 4 |
|  | Trainings for moving objects | 6 |
|  | Trainings for object manipulation | 10 |
|  | Trainings for hand/fingers precision | 4 |
| **Group total** |  | **37** |
| **Lower extremity training** | Training for transfers from sit to stand | 9 |
|  | Training for maintaining standing | 4 |
|  | Training for reaching in standing | 3 |
|  | Training for stepping and walking | 5 |
| **Group total** |  | **21** |
| **Trunk training** | Training for trunk strength in sitting | 4 |
|  | Training for trunk strength in lying position | 3 |
| **Group total** |  | **7** |
| **Training for balance** | Training for balance in sitting | 3 |
|  | Training for balance in standing | 6 |
| **Group total** |  | **9** |
| **Total task training items** |  | **74** |

### Stage 2: Consensus-building to select and refine the tasks to include in the intervention model

Forty-five experts who met the selection criteria were invited. Eighteen panelists consented to participate, of whom 13 (72%) completed the 3 rounds of the Delphi study (Table 3). The Delphi technique took almost a year (January 2023-January 2024) to complete due to delays in response from the pane lists.

**Table 3.** Demographic characteristics of the panelists.

| Panelists (n = 13) |  |
| --- | --- |
| **Gender** |  |
| Male | 6 (46.2%) |
| Female | 7 (53.8%) |
| **Country of residence** |  |
| Nigeria | 7 (53.8%) |
| South Africa | 4 (30.8%) |
| Iran | 1 (7.7%) |
| Thailand | 1 (7.7%) |
| **Experience in stroke rehabilitation** |  |
| Mean number of years | 22 years |

At the end of Delphi round one (S6), 28 (38%) items of the intervention model achieved item validity scores of ≥ 0.78 and were retained. Four of the items received a score of less than ≤ 2 by all the panelists and these items were removed. The remaining 42 items were amended based on the comments and suggestions by the panelists (S5) and re-entered into round two (S7). At the end of the second round, 7 (17%) of the items achieved item validity scores of ≥ 0.78 and were retained.

Thus at the end of the second round, 35 (47%) of the 74 task trainings met the predefined item validity score of ≥ 0.78 and 4 (5%) were removed. The remaining 35 items were considered at the study team meeting. The team (RI, IUL, UMB, AA and one Delphi panelist) agreed on the removal of all 25 items proposed as meriting removal, and 10 items modified as warm up exercises and 4 new task trainings were added, based on the comments and suggestions (S5) made by the panelists in both the previous rounds of the Delphi. Fourteen training tasks were the items added to the already retained 35 items and entered in to round three (S8) for content validation. Figure 1 shows the flow of participants and items through the stages of the study.

**Fig 1.** Flow of the consensus-building process.

At the end of round three a mean content validity score of 0.93 (table 4) was achieved and therefore all the fourteen items from the team meeting were retained.

**Table 4.** The proportion of items giving a rating of 3 or 4 by raters involved in round 3.

| **Rater** | **Score** |
| --- | --- |
| P1 | 0.79 |
| P2 | 1.00 |
| P3 | 0.92 |
| P4 | 0.95 |
| P5 | 0.89 |
| P6 | 0.95 |
| P7 | 0.97 |
| P8 | 0.97 |
| P9 | 0.97 |
| P10 | 0.97 |
| P11 | 0.82 |
| P12 | 0.92 |
| P13 | 1.00 |
| **Mean core** | **0.93** |

The number of items in each category through the stages of the study is shown in Table 5. Thus at the end of round three consensus was reached to retain 49 (66%) task trainings (table 6) of which 10 were retained as warm up exercises (S9).

**Table 5.**
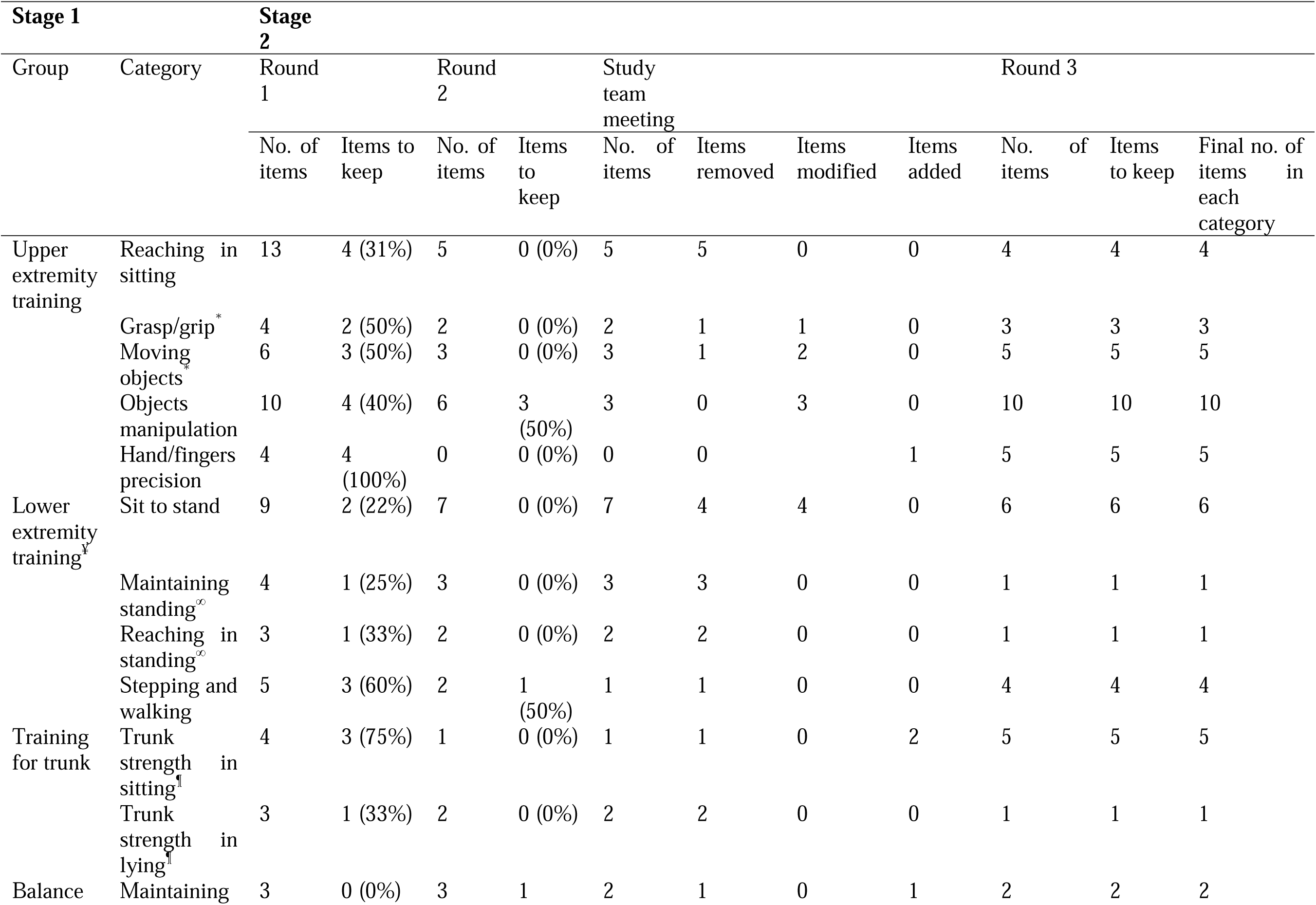
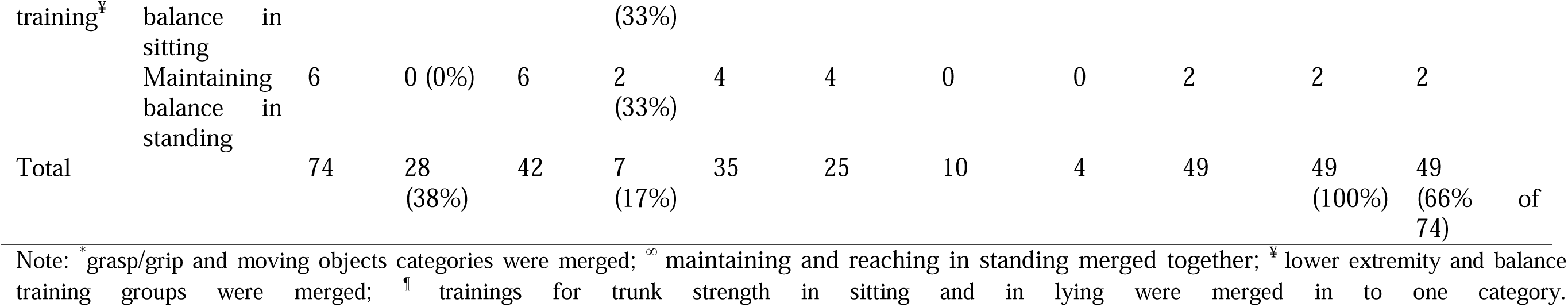
Number (%) of items in each category through the stages of the study.

**Table 6.** Final grouping/categorization of items in the model.

| **Task training group** | **Category** | **No of task training** |
| --- | --- | --- |
| **Upper extremity training** | Warm up | 5 |
|  | Training for reaching | 4 |
|  | Training for grasp/grip and Trainings for moving objects | 8 |
|  | Training for object manipulation | 10 |
|  | Training for hand/fingers precision | 5 |
| **Group total** |  | **32** |
| **Lower extremity and balance training** | Warm up | 2 |
|  | Training for transfers from sit to stand | 2 |
|  | Training for maintaining standing and reaching in standing | 2 |
|  | Training for stepping and walking | 4 |
| **Group total** |  | **10** |
| **Training for trunk strength** | Warm up | 3 |
|  | Training for trunk strength in sitting and lying position | 4 |
| **Group total** |  | **7** |
| **Total task training** |  | **49** |

### Task-specificity of the items

The text responses of the panelists emphasized the importance of making the items of the model to be task-specific training. As a result, items that comprise activities not task-specific received low ratings. The need for warm-up exercises prior to the commencement of the main training was also among the suggestions made by the panelists. In the response to these comments and suggestions, the study team decided to retain those items comprising of non-task-specific activities as warm-up exercises, and this decision was unanimously agreed by the panelists in the final round of the Delphi. Ten items of the model were converted to warm-up exercises: five for upper extremity training group; two for lower extremity and balance training group; and three for the trunk training group.

### Redundant and similar items

The panel highlighted the issue of repetition of items within and outside categories. Redundancy and similarities were the major terms used by the panelists when describing such items, the panelists suggested merging some task trainings together, it was argued and agreed by the panelists that most of the items in the lower extremity training group were similar to those in the balance training group and can be used to improve both lower extremity and balance functions, and consequently the lower extremity and balance training groups were merged as one training group. The following categories were merged based on the panelists’ suggestions: grasp/grip and moving objects; maintaining and reaching in standing; training for trunk strength in sitting and in lying. These merging were undertaken during the study team meeting.

### Non familiar item

Some items involve activities that are peculiar to the culture of the target population and therefore, were not clear to some panelists which prompted them to suggest that “use of pictures for these activities would help because it takes time to figure out what is expected.” Thus, picture demonstration of such activities involving the use of Hausa local cap, eating bowl, and ablution related activities were included in the second round of the Delphi. This resulted in better understanding and facilitated the rating of the items by the panelists.

## Discussion

This article presents a consensus-building study aimed at identifying the relevant and suitable items (task trainings) of a Task-specific Self-Rehabilitation Training (TASSRET) intervention model for improving functional ability outcomes among rural community-dwelling stroke survivors. The outcome is a list of 39 task-specific training and 10 warm up exercises, which can be self-administered at home using common household items that are peculiar to the target population. Due to the limited economic resources of health care systems in developing countries, the emergence of new and expensive technologies has been restricted to developed countries.^4^ The existence of such innovative and inexpensive stroke intervention approaches would greatly benefit stroke rehabilitation particularly in developing countries.^8^

This Task-specific Self-Rehabilitation Training (TASSRET) is a product of combined input from the rural community-dwelling stroke survivors and the expertise of 13 stroke rehabilitation professionals with adequate clinical experience and cumulative learning. The results of this study may augment the poor stroke rehabilitation services as well as promote stroke rehabilitation in rural communities, particularly in the SSA. Rehabilitation is an essential part of universal health coverage and is a key strategy for achieving Sustainable Development Goal 3 – “Ensure healthy lives and promote well-being for all at all ages.”^9^ The TASSRET model may promote inclusiveness and coverage of rehabilitation services by providing the opportunity of self-rehabilitation among rural community-dwelling stroke survivors (including the unlettered) within the comfort of their homes.

This study has some strengths and limitations. The source of the initial list of candidate features for entry into the first round of the Delphi was through focus group discussion with stroke survivors. This is recommended as a valid source of appropriate items to inform the first quantitative round of a Delphi survey.^28^ Several rounds of written detailed information were sent to panelists to ensure that the panelists have an understanding of the study’s aim and process, and this helps build good research relationships between researchers and panelists.^29^ Moreover, reminder emails and sometimes phone calls, were employed to enhance time and rate of response. Nonetheless, the response was at times poor.

A further strength of this study was the administrative skills employed in coding system for tracking panelists and their responses through the Delphi rounds. Analyzing changes of opinion and suggestions was undertaken and managed well. These are issues upon which the smooth execution of a Delphi is based, but are scarcely considered in the literature.^28^

Some of the limitations of this study include the identification of the potential panelists, as the contacts of the potential panelists were obtained through their published papers and in many instances the addresses were obsolete. Also, the study team made an input into the modification of some items prior to the commencement of the final round of the Delphi. Although it was done based on the responses of the panelists, this goes against the basic tenets of the Delphi method. The input from the research team was deemed necessary as it was difficult to progress without doing so. This was similarly noted by Green et al in the Delphi study of general practitioners’ information requirements, in which the study team reordered and reduced the information generated.^30^

Another limitation was in the pursuit of true anonymity. In order to achieve maximum response, the identity of some of the panelists was known to the research team, this helped in persuading panelists that responded late, however, we ensured that the panelists and their responses remained strictly anonymous to the rest of the panel.

## Conclusion

This Task-specific Self-Rehabilitation Training model can be used as an intervention approach to augment and promote stroke rehabilitation among rural community-dwelling stroke survivors, especially in SSA. Further work may be needed to improve and determine the effectiveness of this intervention program in improving functional ability outcomes post-stroke.

## Data Availability

All relevant data are within the manuscript and its Supporting Information files.

## Acknowledgments

We appreciate the National Assembly, Abuja, Bayero University Kano and Skyline University Kano, Nigeria for providing the enabling environment for the writing and submission of this manuscript.

## Supporting information

S1: FGD interview guide

S2: Initial task trainings

S3: Delphi invitation and consent letter

S4: Delphi instruction note

S5: Panelists’ comments and suggestions

S6: Delphi round 1

S7: Delphi round 2

S8: Delphi round 3

S9: Final task trainings (TASSRET)

